# Pediatric Poverty and County-Level Cardiovascular Mortality in the United States: A National Cross-Sectional Analysis

**DOI:** 10.64898/2026.07.15.26358197

**Authors:** Kevin Babapour Digaleh, Mahdi Bouchekouk, Bina Ronen, Ang Sun, Wyatt House, Toshihito Gomibuchi, Alyster Alcudia, G. William Moser, Suyog Mokashi

**Author notes:** Corresponding Author: Suyog Mokashi, MD, MBA 3401 N. Broad Street, Suite 301 Philadelphia, PA 19140.

## Abstract

**Background:** Cardiovascular disease remains the leading cause of death in the United States, and marked geographic disparities in cardiovascular mortality persist. However, the community-level socioeconomic indicators most strongly associated with these disparities remain unclear. Community-level measures capture the social and economic conditions that influence cardiovascular health across populations and may help identify communities at greatest risk. We used the Area Health Resources File (AHRF) to identify socioeconomic measures most strongly associated with county-level cardiovascular mortality.

**Methods:** We performed a national cross-sectional ecological analysis using the 2024–2025 Area Health Resources File (AHRF), including counties in the 50 U.S. states and the District of Columbia. The primary outcome was an AHRF-defined cardiovascular mortality composite derived from 2021–2023 National Center for Health Statistics (NCHS) mortality data. Community-level socioeconomic measures included 2023 overall, pediatric, and family childhood poverty and 2019–2023 overall, female, and White unemployment. County-level associations were evaluated using Spearman rank correlation and regional differences using the Kruskal–Wallis test. Sensitivity analyses used a partial mortality composite and Kendall τ correlation.

**Results:** Among 1,982 counties, cardiovascular mortality varied significantly across U.S. Census divisions (P<0.001), with the highest population-weighted rate in the East South Central division (348.5 deaths/100,000) and the lowest in the Mountain division (233.9 deaths/100,000). Pediatric poverty demonstrated the strongest association with cardiovascular mortality (ρ=0.612), followed by family childhood poverty (ρ=0.603) and overall poverty (ρ=0.524; all P<0.001). In contrast, unemployment measures were more weakly associated (overall ρ=0.209, White ρ=0.176, female ρ=0.141; all P<0.001). Results were consistent in sensitivity analyses.

**Conclusions:** County-level poverty, particularly pediatric poverty, was more strongly associated with cardiovascular mortality than unemployment across U.S. counties. These findings suggest pediatric poverty may serve as a useful community-level indicator for identifying populations at increased cardiovascular risk and prioritizing future public health interventions.

## Introduction

Cardiovascular disease remains one of the leading causes of death in the United States, with substantial geographic variation in cardiovascular mortality across U.S. counties.^1,2^ Persistently high mortality has been documented throughout the South, Appalachia, and the broader Stroke Belt, whereas lower rates are observed in portions of the Mountain West.^2,3^ These disparities have not narrowed uniformly; recent national analyses suggest that differences between high- and low-mortality regions have remained substantial and, in some comparisons, widened over the past two decades.^4,5^ Identifying the community-level factors associated with this persistent geographic variation remains an important public health priority.

A growing body of county-level evidence indicates that socioeconomic conditions and area deprivation are consistently associated with cardiovascular mortality.^6–8^ In recent county-level work, area-based deprivation indices have explained a substantial share of the variation in premature cardiovascular mortality, and changes in county economic prosperity have tracked changes in cardiovascular mortality among middle-aged adults over time.^7,9^ State-of-the-art reviews of the social determinants of cardiovascular health identify economic stability, including income and material resources, as an important social determinant of cardiovascular health.^6,10^ Poverty, in this context, can be understood as a measure of chronic material deprivation that may reflect upstream constraints on access to preventive care, food and housing security, educational and economic opportunity, and exposure to chronic psychosocial stress.

Individual-level studies consistently link childhood socioeconomic disadvantage with increased adult cardiovascular risk through life-course biological, behavioral, and social mechanisms.^11–15^ These observations provide a rationale for evaluating pediatric poverty as a community-level marker of broader socioeconomic disadvantage, although ecological associations cannot establish individual-level causation. Whether pediatric poverty is more strongly associated with county-level cardiovascular mortality than other readily available measures of poverty and unemployment remains unclear.

In this national, county-level analysis we therefore had two aims: first, to characterize contemporary geographic variation in an AHRF-defined cardiovascular mortality composite across U.S. counties, Census divisions, and states; and second, to compare the county-level associations between cardiovascular mortality and three poverty measures (overall, pediatric, and family childhood poverty) and three unemployment measures (overall, female, and White). Particularly, pediatric poverty was evaluated as a community-level correlate of cardiovascular mortality rather than as an individual-level causal exposure.

## Methods

### Data source

A retrospective cross-sectional analysis using the 2024–2025 release of the Area Health Resources File (AHRF), a publicly available county-level dataset maintained by the U.S. Health Resources and Services Administration, was performed.^16^ Within AHRF, cause-specific mortality counts originate from the National Center for Health Statistics (NCHS) Mortality Detail Data Files; population denominators from the U.S. Census Bureau Vintage 2023 County Population Estimates; poverty estimates from the 2023 Small Area Income and Poverty Estimates (SAIPE) program; and unemployment estimates from the American Community Survey (ACS) 2019–2023 5-year tables.

### Study population and geographic units

The study population comprised all counties and county-equivalents in the 50 U.S. states and the District of Columbia (n=3,153); U.S. territories were excluded. Connecticut was excluded from county-level analyses because incompatible county definitions across AHRF data sources precluded linkage of mortality, population, and socioeconomic data.

### Cardiovascular mortality outcome

The primary outcome was an AHRF-defined cardiovascular mortality composite calculated as the sum of 2021–2023 average annual deaths from ischemic heart disease (ICD-10 I20–I25), cerebrovascular disease (I60–I69), and other cardiovascular disease (I00–I09, I10–I15, I26–I51, I70, I71–I78). The strict primary composite required all three components to be available for a county; a partial lower-bound composite including counties with at least one observed component was used for sensitivity analysis. Crude rates per 100,000 residents were calculated using 2023 Census county population estimates. Because AHRF does not provide age-adjusted cause-specific rates or formally define this summed measure as total cardiovascular mortality, we refer to it as an AHRF-defined cardiovascular mortality composite.

### Socioeconomic predictors

Community-level socioeconomic predictors included three 2023 Small Area Income and Poverty Estimates (SAIPE) measures: overall poverty, pediatric poverty (ages 0–17), and family childhood poverty (related children ages 5–17). Three unemployment measures were derived from the 2019–2023 American Community Survey (ACS): overall, female, and White unemployment among the civilian labor force aged ≥16 years. ACS unemployment estimates represent pooled 5-year averages.

### Geographic aggregation

State- and Census division–level cardiovascular mortality rates were calculated as population-weighted crude rates (per 100,000) by aggregating county deaths and populations, ensuring estimates reflected population size rather than unweighted county averages.

### Statistical analysis

County-level associations between cardiovascular mortality and socioeconomic predictors were evaluated using Spearman rank correlation. Differences in county-level cardiovascular mortality across the nine U.S. Census divisions were assessed using the Kruskal–Wallis test. State-level comparisons were reported descriptively because differential suppression limited comparable county representation. Statistical significance was defined as P<0.05 (two-sided). Analyses were performed in Python 3 using pandas and scipy.stats.

### Sensitivity analyses

Sensitivity analyses repeated all correlations and geographic summaries using the partial lower-bound cardiovascular mortality composite and re-estimated county-level correlations using Kendall τ.

## Results

### Analytic sample

Among 3,153 U.S. counties and county-equivalents, 1,982 contributed to the primary AHRF-defined cardiovascular mortality composite. The partial lower-bound sensitivity analysis included 2,765 counties.

### Geographic variation in cardiovascular mortality

Cardiovascular mortality differed significantly across the nine U.S. Census divisions (*P*<0.001). The East South Central division had the highest population-weighted cardiovascular mortality rate (348.5 deaths/100,000), whereas the Mountain division had the lowest (233.9 deaths/100,000) (Figure 2).

State-level cardiovascular mortality was highest in West Virginia (392.8 deaths/100,000), Mississippi (385.7), and Alabama (377.2), and lowest in Utah (160.7), Alaska (176.0), and Colorado (188.0).

### Socioeconomic correlates of cardiovascular mortality

All socioeconomic measures were significantly associated with cardiovascular mortality (*P*<0.001). Poverty measures demonstrated substantially stronger associations than unemployment measures. Pediatric poverty showed the strongest correlation (ρ=0.612), followed by family childhood poverty (ρ=0.603) and overall poverty (ρ=0.524). Among unemployment measures, overall unemployment demonstrated the strongest association (ρ=0.209), followed by White unemployment (ρ=0.176) and female unemployment (ρ=0.141). Notably, even the weakest poverty measure correlated more strongly with cardiovascular mortality than the strongest unemployment measure (Figure 1; Table 2).

**Figure 1.**
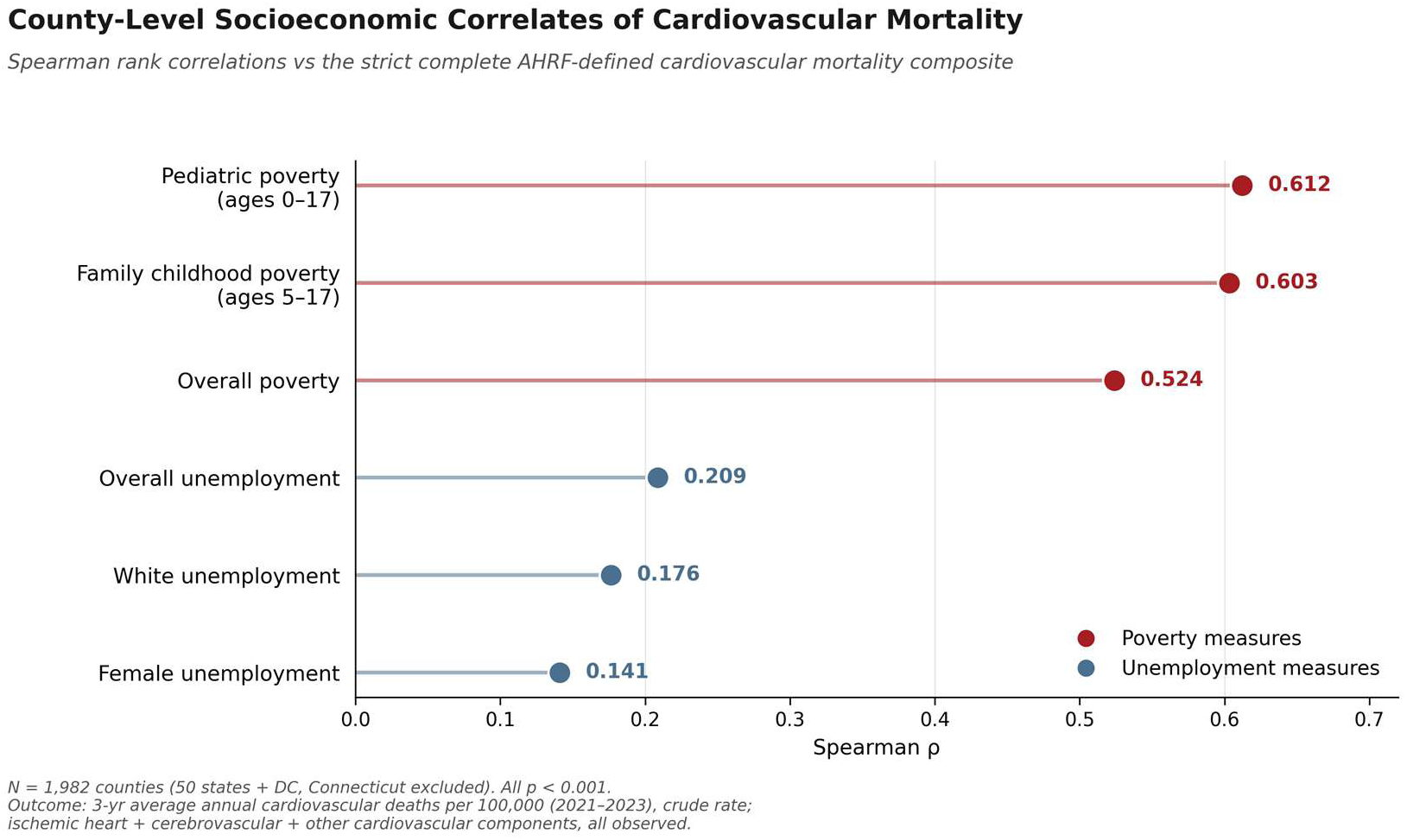
**County-Level Socioeconomic Correlates of Cardiovascular Mortality** Spearman rank correlations between county-level socioeconomic measures and the AHRF-defined cardiovascular mortality composite. Poverty measures (red)—pediatric poverty (ages 0–17), family childhood poverty (ages 5–17), and overall poverty—demonstrated stronger associations with cardiovascular mortality than unemployment measures (blue)—overall, White, and female unemployment. All correlations were statistically significant (*P*<0.001). The analysis included 1,982 counties; Connecticut was excluded because incompatible geographic definitions prevented county-level linkage of mortality and socioeconomic data.

**Figure 2.**
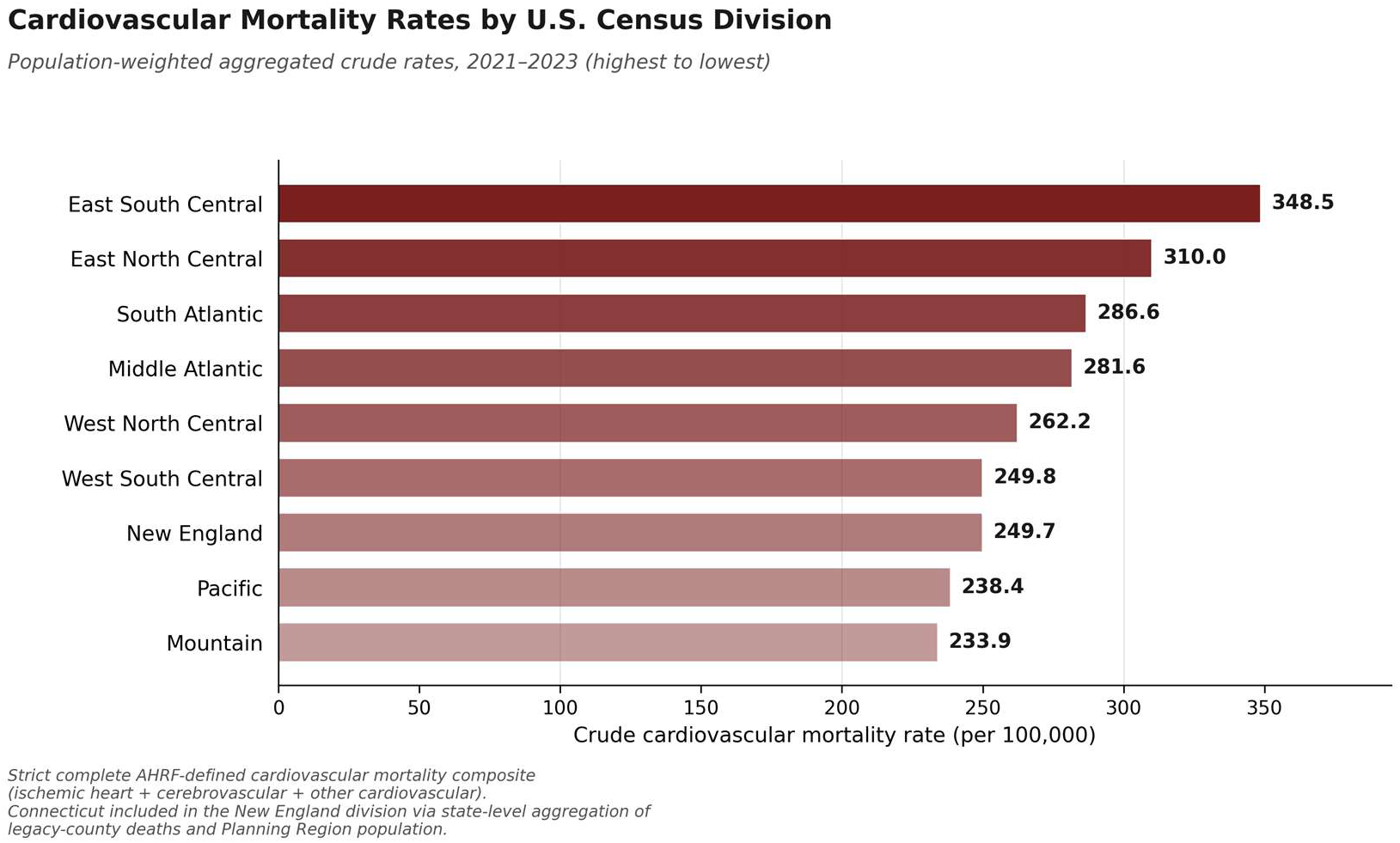
**Cardiovascular Mortality Rates by U.S. Census Division** Population-weighted crude cardiovascular mortality rates (per 100,000 residents) across the nine U.S. Census divisions using the primary AHRF-defined cardiovascular mortality composite. The East South Central division had the highest mortality rate (348.5 per 100,000) and the Mountain division the lowest (233.9 per 100,000). County-level differences were significant (Kruskal–Wallis *P*<0.001).

**Table 1.** Analytic sample and county-level measures in the AHRF cardiovascular mortality analysis Domain / measure Source and time period Operational definition or analytic.

| Domain / measure | Source and time period | Operational definition or analytic sample |
| --- | --- | --- |
| ***Panel A. Analytic sample*** |  |  |
| Geographic universe | U.S. Census Bureau geography | 50 states and the District of Columbia; 3,153 counties/county-equivalents after exclusion of U.S. territories |
| County-level Connecticut handling | AHRF 2024–2025 release | Connecticut excluded from county-level analyses<sup>a</sup> |
| Strict primary analytic sample | Counties contributing to the strict complete-component composite | n=1,982 counties |
| Partial lower-bound sensitivity sample | Counties contributing to the partial lower-bound composite | n=2,765 counties |
| ***Panel B. Cardiovascular mortality outcomes*** |  |  |
| Strict complete-component AHRF-defined cardiovascular mortality composite<sup>b</sup> (primary) | NCHS Mortality Detail Data Files (in AHRF); 2021–2023 3-year average annual deaths | Sum of three component categories<sup>c</sup>; defined only when all three components are observed for a county. Crude rate per 100,000<sup>d</sup> using 2023 Census County Population Estimates as denominator |
| Partial lower-bound composite (sensitivity) | NCHS Mortality Detail Data Files (in AHRF); 2021–2023 3-year average annual deaths | Sum of observed components when at least one is observed; lower-bound interpretation in counties with partial NCHS suppression. Same denominator and rate scale as primary outcome |
| ***Panel C. Socioeconomic predictors*** |  |  |
| Overall poverty | SAIPE; 2023 single-year estimate | Percent of all persons in poverty |
| Pediatric poverty (ages 0–17) | SAIPE; 2023 single-year estimate | Percent of persons aged 0–17 in poverty |
| Family childhood poverty (ages 5–17) | SAIPE; 2023 single-year estimate | Percent of related children aged 5–17 in families in poverty |
| Overall unemployment | ACS 2019–2023 5-year estimate (Table B23001) | Unemployed persons aged ≥16 as a percent of the civilian labor force aged ≥16 |
| Female unemployment | ACS 2019–2023 5-year estimate (Table B23001) | Unemployed females aged $\geq 16$ as a percent of the female civilian labor force aged $\geq 16$ |
| White unemployment<sup>e</sup> | ACS 2019–2023 5-year estimate (Table C23002A) | Unemployed White persons aged $\geq 16$ as a percent of the White civilian labor force aged $\geq 16$ |

*AHRF = HRSA Area Health Resources File (2024–2025 release); NCHS = National Center for Health Statistics; ICD-10 = International Classification of Diseases, 10th Revision; SAIPE = U.S. Census Bureau Small Area Income and Poverty Estimates; ACS = American Community Survey.*
<sup>a</sup> Connecticut was excluded from county-level analyses because, following the state’s 2022 transition from legacy counties to Planning Regions, no Connecticut row contains both a mortality numerator and the population and predictor data required to calculate matched county-level outcomes. Connecticut was retained in state- and Census-division-level analyses via aggregation of mortality across the 8 legacy counties and population across the 9 Planning Regions.
<sup>b</sup> Described throughout as an “AHRF-defined cardiovascular mortality composite” because HRSA does not formally label the sum of these three NCHS categories as a unified construct, and a small number of ICD-10 circulatory codes are not represented in the AHRF cardiovascular block.
<sup>c</sup> Component categories: ischemic heart disease (ICD-10 I20–I25); cerebrovascular disease (I60–I69); and other cardiovascular disease (I00–I09, I10–I15, I26–I51, I70, I71–I78).
<sup>d</sup> All reported rates are crude. AHRF does not provide age-adjusted cause-specific cardiovascular mortality measures.
<sup>e</sup> ACS White unemployment is constructed from ACS Table C23002A and includes both Hispanic and non-Hispanic White persons.

**Table 2.** County-level associations between socioeconomic measures and the AHRF-defined cardiovascular mortality composite.

| Predictor | Strict<br>Spearman $\rho$ | P value | Partial<br>Spearman $\rho$ | P value | Strict Kendall $\tau$ | P value |
| --- | --- | --- | --- | --- | --- | --- |
| Pediatric poverty (ages 0–17) | 0.612 | <0.001 | 0.535 | <0.001 | 0.437 | <0.001 |
| Family childhood poverty (ages 5–17) | 0.603 | <0.001 | 0.526 | <0.001 | 0.431 | <0.001 |
| Overall poverty | 0.524 | <0.001 | 0.461 | <0.001 | 0.370 | <0.001 |
| Overall unemployment | 0.209 | <0.001 | 0.226 | <0.001 | 0.141 | <0.001 |
| White unemployment | 0.176 | <0.001 | 0.196 | <0.001 | 0.120 | <0.001 |
| Female unemployment | 0.141 | <0.001 | 0.166 | <0.001 | 0.094 | <0.001 |

$\rho$ = Spearman rank correlation coefficient; $\tau$ = Kendall rank correlation coefficient. Pairwise complete-case sample sizes are identical across the six predictors within each outcome (n=1,982 for the strict composite; n=2,765 for the partial lower-bound composite). Connecticut was excluded from county-level analyses because mortality and population/predictor data were available for nonmatching county-equivalent geographies. All tests were two-sided.

### Sensitivity analyses

Results were robust across sensitivity analyses. The partial lower-bound composite yielded the same ranking of socioeconomic predictors, with pediatric poverty remaining the strongest correlate of cardiovascular mortality. Geographic patterns were likewise unchanged, with the East South Central and Mountain divisions retaining the highest and lowest mortality rates, respectively. Similar findings were observed using Kendall τ correlation analysis.

## Discussion

In this national county-level analysis, the AHRF-defined cardiovascular mortality composite demonstrated marked geographic variation across the United States, with the highest population-weighted mortality in the East South Central Census division and the lowest in the Mountain division. Among six community-level socioeconomic measures, all three poverty measures were more strongly associated with cardiovascular mortality than unemployment measures, with pediatric poverty (ages 0–17) emerging as the strongest correlate. These findings identify community-level socioeconomic patterns associated with cardiovascular mortality across U.S. counties.

The geographic distribution of cardiovascular mortality observed in this study is consistent with prior county-level analyses demonstrating persistently elevated mortality throughout the South, Appalachia, and the broader Stroke Belt, with lower mortality in the Mountain West.^2–5^ Although differences in outcome definitions and analytic methods preclude direct comparison of rankings, our findings provide a contemporary national assessment using 2021–2023 mortality data and reinforce the persistence of substantial regional disparities in cardiovascular mortality.

Socioeconomic deprivation is a well-established determinant of cardiovascular health, and previous county-level studies have shown that area deprivation explains a substantial proportion of geographic variation in cardiovascular mortality.^6–10^ Consistent with this literature, poverty measures were substantially more strongly associated with cardiovascular mortality than unemployment measures. Poverty likely reflects broader and more sustained material deprivation, encompassing limited access to preventive care, housing and food insecurity, educational opportunity, chronic psychosocial stress, and other social determinants that influence cardiovascular health across populations.^6–10^

Pediatric poverty demonstrated the strongest association with cardiovascular mortality. Individual-level studies consistently associate childhood socioeconomic disadvantage with increased adult cardiovascular risk through biological, behavioral, and social pathways, supporting a life-course relationship between early socioeconomic conditions and later cardiovascular disease.^11–15^ However, our analysis cannot determine whether childhood poverty directly contributed to cardiovascular mortality among the individuals represented in the mortality data. Rather, pediatric poverty should be interpreted as a community-level marker of sustained socioeconomic disadvantage that may identify counties at particularly high cardiovascular risk.

The weaker associations observed for unemployment should not be interpreted as evidence that unemployment is unimportant for cardiovascular health.^10^ Rather, unemployment represents a narrower and more transient measure of economic conditions, whereas poverty better captures long-term material deprivation and accumulated socioeconomic disadvantage. Consequently, poverty measures may provide more informative community-level indicators of cardiovascular mortality risk in cross-sectional analyses.

These findings suggest that readily available community-level poverty measures, particularly pediatric poverty, may help identify communities with the greatest cardiovascular mortality burden and inform population health surveillance, prevention efforts, and resource allocation.^6,11^ Future studies should evaluate whether incorporating pediatric poverty into community risk assessment improves targeting of cardiovascular prevention strategies.

This study has several limitations. First, the ecological cross-sectional design precludes causal inference and individual-level interpretation. Second, AHRF provides only crude county-level cardiovascular mortality rates, preventing age adjustment. Third, suppression of NCHS mortality counts excluded many rural and low-population counties from the primary analysis, although sensitivity analyses produced consistent findings. Fourth, Connecticut was excluded from county-level analyses because incompatible geographic definitions across AHRF data sources prevented linkage of mortality and socioeconomic data.

Finally, modest temporal differences between mortality, poverty, and unemployment data and the potential for residual confounding are inherent limitations of ecological analyses.

In conclusion, county-level poverty, particularly pediatric poverty, was more strongly associated with cardiovascular mortality than unemployment across U.S. counties. These findings identify pediatric poverty as a readily available community-level marker of cardiovascular mortality burden that warrants further investigation and may help inform population health surveillance and cardiovascular prevention efforts.

## Data Availability

The data analyzed in this study are publicly available from the respective data providers and can be accessed through their official websites.

## Acknowledgments

None

## Institutional Review Board Statement / Ethical Considerations

Ethics determination to be finalized. The Area Health Resources File is a publicly available, aggregated, de-identified county-level dataset; the study is expected to qualify for IRB exemption or non-human-subjects determination pending formal institutional confirmation.

## Data Availability

The 2024–2025 Area Health Resources File used in this analysis is publicly available from the U.S. Health Resources and Services Administration at https://data.hrsa.gov/topics/health-workforce/ahrf. Underlying county-level cause-of-death counts are available from the CDC WONDER platform (https://wonder.cdc.gov). Analytic code and derived intermediate datasets supporting the results reported here are available from the corresponding author upon reasonable request.

## Common notes (apply to both figures)

- Source: HRSA Area Health Resources File (AHRF) 2024–2025 release, incorporating National Center for Health Statistics mortality data, U.S. Census Bureau County Population Estimates and Small Area Income and Poverty Estimates (SAIPE), and the American Community Survey 2019– 2023.
- The cardiovascular mortality outcome is an AHRF-defined cardiovascular mortality composite derived from ischemic heart disease, cerebrovascular disease, and other cardiovascular disease mortality.
- Mortality rates are crude (not age-adjusted) because age-adjusted cause-specific rates are not available in the AHRF.
- Sensitivity analyses using the partial lower-bound cardiovascular mortality composite produced similar findings.

## Notes

### Competing Interest Statement

The authors have declared no competing interest.

### Clinical Trial

N/A

### Author Declarations

Institutional Review Board (IRB) approval was not required because this study used publicly available, de-identified data and did not involve human subjects research as defined under applicable federal regulations.

